# Risk Communication and Public Health Emergency Responses during COVID - 19 Pandemic in Rural Communities in Kenya: A Cross-sectional Study

**DOI:** 10.1101/2024.11.15.24317380

**Authors:** Wilberforce Cholo, Fletcher Njororai, Walter Ogutu Amulla, Caleb Kogutu Nyaranga

**Affiliations:** Department of Public Health, Masinde Muliro University of Science and Technology; 190-50100, Kakamega, Kenya; Department of Public Health, The University of Texas at Tyler, Tyler, TX 75799, USA.; Department of Public Health, Kisii University; 408-40200, Kisii, Kenya; Department of Environmental and Occupational Health, Kenyatta University, Nairobi 43844-00100, Kenya

**Keywords:** Risk communication, Perception, COVID-19, Vaccine acceptance, Emergency, credibility, Public Health

## Abstract

**Background:** COVID-19 pandemic highlighted the crucial role of community preventive behaviors in controlling the virus’s spread. Studies show that people’s risk perceptions and awareness significantly contribute to containment and prevention of infections. However, limited studies focused on the influence of risk communication on Public Health Emergency Responses during g the peak of the COVID-19 pandemic in Kenya. This study aimed at assessing the role of risk communication on Public Health Emergency Responses during COVID - 19 Pandemic during the COVID-19 pandemic rural communities in Kenya.

**Methods:** A descriptive cross-sectional study was conducted using a quantitative research approach, collecting data from 806 individuals across Kisumu, Vihiga, and Kakamega counties. Descriptive statistics were used to detail the demographic characteristics of the study population, while logistic regression analysis estimated the associations between risk communication and demographic characteristics on COVID-19 vaccine acceptance, compliance with mitigation behaviors, perceived severity, and perceived susceptibility.

**Results:** The results showed that 55% of participants were male, and 45% were female, with an average moderate compliance with safety measures (Mean = 5.15). A significant portion of participants wore face masks (85.3%), practiced hand hygiene (78.9%), and avoided close contact behaviors (66.6%). Most respondents received information through mass media (86.1%) and health workers (72.9%). Compliance with COVID-19 mitigation measures was highest among those who trusted information from official institutions, health professionals, and mass media, compared to social media, with increased odds of 2.7 times and 2.5 times, respectively. Higher risk perception was significantly associated with older age groups (above 50 years), being male, and working in the private sector. Risk communication significantly influenced risk perception, compliance with COVID-19 measures, and vaccination.

**Conclusion:** The findings suggest that effective risk communication strategies are essential during public health emergencies hence implications for future public health crises. The results underscore the importance of targeted communication and tailored interventions to improve compliance and vaccine acceptance among different demographic groups, ensuring a more robust public health response during outbreaks.

## Background

The COVID-19 pandemic, which nearly halted global activities in 2020 and has claimed over 7 million lives worldwide [1], highlights the critical need for enhanced international cooperation, preparedness, global security, surveillance, monitoring, and capacity building for public health emergencies (PHEs) and risk communication [1,2]. The International Monetary Fund estimates that the pandemic will cost the global economy approximately US$13.8 trillion by the end of 2024 [3], in addition to its significant socioeconomic and other broad societal impacts. It is widely regarded as the most severe disaster in living memory by nearly any measure [4]. With the increasing frequency of PHEs, the World Health Organization (WHO) emphasizes the importance of risk communication as one of the major areas of focus for global investment, collaboration, research, data sharing, and policy development [5–7].

In 2015, following the Ebola Virus Disease crisis in West Africa, WHO established a working group to draft guidelines on building national capacities for communicating health risks during PHEs [8]. Risk communication has been identified as one of the eight core capacities under the International Health Regulations (IHR of 2005) [9]. There is a global agenda to advance breakthroughs in risk and crisis communication to support evidence-based campaigns in PHEs [5].

Public health emergencies encompass many happenings which include disease outbreaks (epidemics and pandemics), environmental disasters, humanitarian crises, and other man-made disasters which may be in localized geographical locations or widespread in large areas or across countries and the world in general [10]. Large-scale epidemics and pandemics cause widespread death and suffering, disproportionately affecting vulnerable populations, and lead to extensive social, economic, health, and political disruptions, complicating recovery efforts [7]. Therefore, timely and effective responses are critical but challenging.

During PHEs, it is essential for people to understand the health risks they face, the nature and sources of these risks, their potential impact at individual and community levels, and the actions they should take to protect their health and that of their communities [8]. Risk communication (RC) involves the real-time exchange of information, advice, and opinions among experts, community leaders, officials, and the public at risk [11]. Effective RC aims to promote health behaviors, such as adherence to public health and social measures (PHSM), timely screening, and seeking prompt treatment and other mitigation measures [12]. Before the widespread availability of biomedical innovations like COVID-19 tests, vaccines, and treatments, PHSM were crucial in limiting the virus’s spread [13]. PHSM help frame risks and risk perception and are integral to managing PHEs beyond just biomedical interventions [12]. Risk perceptions during PHEs, and related actions and responses vary and evolve as the situation and circumstances making imperative an iterative process for effective RC.

Effective RC influences how people perceive risks and prompts desired actions during emergencies. It is a dynamic, interactive, and adaptive process [8, 11, 18]. However, risk communication models developed in Western countries often fail to resonate with African communities due to contextual differences in risk perception, stereotypes, and other factors like wars, displacement, health literacy, negative colonial medical legacy and practices, and weak health care infrastructure [4, 19]. Following WHO’s pandemic declaration in March 2020, African countries quickly implemented countermeasures to curb COVID-19’s spread to mitigate adverse impacts such as increasing global morbidity, mortality, and overwhelmed health systems [20, 21]. The WHO Regional Office for Africa’s risk communication and community engagement (RCCE) framework guided many African countries’ responses, with varied outcomes [23]. In May 2022, the Africa Centers for Disease Control and Prevention (Africa CDC), in collaboration with WHO, USAID, UNICEF, and other partners, established the Public Health Risk Communication and Community Engagement of Practice for Africa (PH-RCCE-CoPA) [22]. Despite these efforts, some African countries faced challenges such as resource shortages, staff constraints, and weak coordination strategies [23].

A study on RCCE strategies for COVID-19 in 13 African countries, including Kenya, identified key RCCE strategies, including risk communication systems, community engagement, public communication, and managing misinformation [24]. Challenges included government distrust, cultural resistance, and misinformation. A case study of Kenya’s early COVID-19 response (February 2020 – May 2021) found initial significant compliance with non-pharmaceutical measures such as PHSMs due to strict governmental enforcement but that shifted to a significant subsequent decline in compliance once lockdowns were eased, as the perceived risk in people and communities diminished [25]. However, limited studies focused on the influence of risk communication on risk perception and compliance with preventive behaviors during the peak of the COVID-19 pandemic in Kenya. This study aimed at assessing the role of risk Communication and public health emergency responses during COVID - 19 pandemic in rural communities in Kenya.

## Materials and Methods

### Research Design

A cross-sectional design using quantitative research strategy was executed to assess the role of risk Communication and factors influencing public health emergency responses during COVID - 19 pandemic in selected rural communities in Kenya. The study was conducted in May-August 2021, at the peak of COVID-19 globally.

### Study setting

The study was carried out in Kakamega, Vihiga, and Kisumu Counties in Western Kenya. Kakamega County covers an area of approximately 3,050.3 km². The county has twelve sub-counties, eighty-three locations, two hundred and fifty sub-locations, one hundred eighty-seven Village Units and four hundred Community Administrative Areas. There are 433,207 households with an average size of 4.3 persons per household, a population of 1,867,579, and a population density of 618 people per square.

Vihiga County lies in the Lake Victoria Basin and covers an area of 531.0 Km2. Vihiga County is located around 80 km northwest of Eldoret, around 60 km north of Kisumu, and approximately 350km west of Nairobi City, the capital city of Kenya. It has a population of 590,013 of which 51.9% are females while male constitutes 48.1%. Sixty-four-point four percent (64.4%) of the total population are under the age of 30 [26]. The County has five administrative Sub-Counties. The county is further subdivided into 38 locations, and 131 sub-locations.

Kisumu County is bordered to the north by Nandi County and to the north east by Kericho County. The land area of Kisumu County totals 2085.9 km^2.^ [26]. It has a population of 1,155,574 of which women make up 50.1% of Kisumu’s population and men represent 49.9%. Sixty-four percent of the total population are under the age of 25 [27]. The land area of Kisumu County totals 2085.9 km^2^ (administratively, the county is divided into 7 sub-counties, and these are further divided into 35 wards [27].

### Study population and participants

All community members aged 18 and above were eligible for participation in the study.

### Sample size determination

An a priori power analysis was conducted using the **G*Power** calculator to determine the required sample size for the study. The analysis was based on the following parameters:

- **Effect size**: 10% of the population,
- **Power**: 80% (to detect meaningful differences),
- **Significance level**: 5% (α = 0.05).

The power analysis for a one-sample proportion test indicated that a minimum sample size of **779 participants** was needed to detect statistically significant effects. To account for a 10% non-response rate, an additional 78 participants were added, bringing the total sample size to **857**. This sample size is sufficient to address the research questions, detect statistical differences, and enhance the generalizability of the findings. The large sample also helps minimize sampling error and ensures the study has adequate statistical power (28).

### Sampling procedure

The 3 counties in Western Kenya were selected using purposive sampling because they were among those in the top 15 leading in highest COVID-19 prevalence in Kenya and the top three leading in COVID - 19 prevalence in Western region of Kenya [29]. Proportionate stratified sampling method was used to select two sub-counties from each selected county based on whether urban or rural and to select the study subjects from the six sub-counties. Two Wards per sub-county were selected using simple random sampling from each selected sub county. A list of households was generated based on administrative location headed by the Chief in the selected sub-counties. Systematic random sampling was then used to select households in the selected Wards. A representative of the eligible study subjects or house heads in the selected households were randomly picked to participate in the study (see Table 1 below).

**Table 1:** Sampling frame.

| County | Sampled sub-county | Sub-county population | Wards(population) | Sample | Total Sampled individuals |
| --- | --- | --- | --- | --- | --- |
| Kakamega | Kakamega Central | 188,212 | Butsotso central (25744) | 149 | 219 |
|  |  |  | Mahiakalu (12067) | 70 |  |
|  | Navakholo | 137165, | Bunyala Central(38407) | 142 | 187 |
|  |  |  | Ingotse-Matiha(12091) | 45 |  |
| Kisumu | Kisumu Central | 174,145 | Kondele (48004) | 93 | 130 |
|  |  |  | Milimani(18902) | 37 |  |
|  | Nyando | 161,508 | Ahero(31440) | 60 | 121 |
|  |  |  | Onjiko(30937) | 51 |  |
| Vihiga | Emuhaya | 69,250 | North East Bunyore(35908) | 36 | 64 |
|  |  |  | Central Bunyore(27316) | 28 |  |
|  | Hamisi | 148 259 | Banja (22535) | 45 | 85 |
|  |  |  | Tambua(18689) | 40 |  |
| Total |  |  |  |  | 806 |

### Variables

#### Dependent variables

The dependent variable for this study was public health emergency responses measured in various ways such as adherence to the public health guidelines and mitigation measures, risk perception and vaccine acceptance

*Compliance behaviors*: hand washing, face mask wearing, sanitization, social distancing, cough hygiene, not touching face, and compliance with other WHO guidelines (including lock down and travel policies). Participants were asked for their compliance with these behaviors and their responses were categorized as: not at all, rarely, frequently and always. The level of compliance was classified as high and low.

*Vaccine acceptance*—defined as the degree to which individuals accept, question, or refuse vaccination. It is one of the major determinants of vaccine uptake rate, vaccine hesitancy, and consequently vaccine distribution success.

*Risk perception*- is the subjective judgement that people make about the characteristics and severity of a risk-focused on risk severity and risk susceptibility. This was classified as either low or high

#### Independent variables

*Risk communication sources* - mass media, social media (X, Facebook and others), various institutions including official health institutions and agencies, and health professionals, and various websites (WHO, CDC, government websites, and more).

*Socio-demographic characteristics* such age, gender, level of education, county of residence, area of residence, occupation, income level and compliance behaviors of respondents. The study focused on six.

#### Data sources and instrument

Data was gathered using a structured questionnaire. The questions were adopted and modified from similar studies [30, 33]. In addition, the WHO’s and Kenyan Government Ministry of Health guidelines on COVID-19 infection prevention and control (IPC) were reviewed and used to refine the questionnaire [34, 36]. The instrument was designed in English and translated into local languages (Luo and Luhya) and was back translated to check for consistency. The instrument had four sections. The study’s questionnaire was divided into three sections. The first section collected sociodemographic data (age, sex, county, education, and income) and assessed individual and community compliance with COVID-19 containment measures and sources of communication and the trusted sources of communication. The second section focused on COVID-19 vaccine acceptance and hesitancy, asking participants whether they or their family members would receive the vaccine when available. Those answering “no” or “unsure” were classified as hesitant. The third section, based on the Health Belief Model, explored predictors of vaccine acceptance through questions on perceived susceptibility, severity, benefits, and cues to action, using a four-point Likert scale.

A pilot study was carried out with 80 people who were chosen from the selected sub-counties but were omitted from the actual study. This was to assess the consistency, clarity, and accuracy, for making any necessary adjustments to refine the study instrument to reduce information bias.

#### Data collection Procedure

Ethical and administrative approval and permission were sought and obtained from the administrative offices of the selected sub-counties in the study. The approval and permission to conduct the study were obtained from the University of Eastern Africa, Baraton Institutional Research Ethics Committee (IREC), and the National Commission for Science, Technology and Innovation (NACOSTI) in Nairobi, respectively. Informed consent was sought and obtained from the study participants. The informed consent ensured that the study adhered to ethical considerations in how it was conducted. The study participants were asked about their willingness to participate in the study after being given information on the purpose and procedures involved in the study. The respondents were given all the relevant information about the study to be undertaken to allow for voluntary consent without coercion, pressure, or undue enticement. Those who accepted to participate signed an informed consent form. The participants were also informed that their participation in the study was voluntary, and anyone could withdraw from the study at any time without repercussions or any penalty. Six trained research assistants collected the study data for a period of two months from July to August 2021 after the second COVID-19 wave in Kenya. The structured questionnaire was self-administered but those who were unable to read and write were assisted by the research assistants.

#### Data Analysis

The collected quantitative data were cleaned, coded, and entered in SPSS version 26 program. Data was cleaned and checked for any errors in data entry. Data analysis was performed using the descriptive statistics and inferential statistics. Descriptive statistics were used to present the demographic characteristics of the study population and were presented as frequencies, proportions, means and standard deviations. Logistic regression analysis was used to estimate the association (crude odds ratio) between COVID-19 acceptances, compliance to COVID-19 mitigation behaviors, perceived severity and perceived susceptibility and demographics and the risk communication (*p*-value < 0.05). *P-value* ≤ 5% is considered statistically significant at 95% CI.

All covariates were added to the model simultaneously. The significance level was set at 0.05. Data were presented in tables, narrative report and a graph.

## RESULTS

Out of the 857 randomly selected participants, 806 (94.2%) consented and completed the questionnaire. In this analysis, the focus was on participants with data on all relevant study variables related to risk perception and public health emergency responses and the sample size was n=806 (100%) participants in the study. These participants had no missing information on key demographic data communication, risk perception, compliance behaviors and vaccine acceptance.

### Demographic Characteristics of the respondents

As shown in Table 2 below, most study participants were males (55.0%), aged 18 to 30 years old (37.8%), and possessed secondary level of education (42.9%). More participants (57%) lived in rural areas and slightly over half (50.5%) of the participants were employed. The mean age of the participants was 35.9 (SD= 13.07) and about 70% of the participants were in the age range of 18 - 40 years, while the modal age group was 18-30 years. Slightly over half (52.6%) of the participants resided in Kakamega County followed by 33.9% who resided in Kisumu County while 13.4% of the participants were from Vihiga County.

**Table 2:** Demographic Characteristics of the respondents.

| **Variable** | **Frequency (No.)** | **Percentage (%)** |
| --- | --- | --- |
| *Gender* |  |  |
| Male | 442 | 55 |
| Female | 364 | 45 |
| *Age* |  |  |
| 18-30 | 305 | 37.8 |
| 31-40 | 264 | 32.8 |
| 41-50 | 96 | 11.9 |
| 51-60 | 76 | 9.4 |
| 61-70 | 45 | 5.5 |
| >70 | 20 | 2.5 |
| *Education level* |  |  |
| No Education | 67 | 8.0 |
| Primary | 106 | 13.2 |
| Secondary | 335 | 42.9 |
| University | 298 | 36.9 |
| *County of residence* |  |  |
| Kakamega | 406 | 50.4 |
| Kisumu | 251 | 31.1 |
| Vihiga | 149 | 18.5 |
| *Area of residence* |  |  |
| Rural | 461 | 57.3 |
| Urban | 345 | 42.8 |
| *Occupation* |  |  |
| Civil Servant | 113 | 14 |
| Private Sector | 96 | 11.9 |
| Self Employed | 190 | 23.6 |
| Non-Employed | 407 | 50.5 |

### Compliance with safety measures and vaccine acceptance

Compliance with WHO safety measures, on average, the participants complied with safety measures (Mean = 5.15). Majority of the participants wore face mask (85.3%), practiced hand hygiene (78.9%) avoided hand shaking, hugging and kissing (66.6%), and practiced cough hygiene (66.0%). Compliance with other WHO COVID-19 guidelines consisting of travel and lock down policies accounted for 62.7%. Social distancing was least adopted (60.2%). Overall, vaccine acceptance rate was low at 40%.

### Risk communication

Table 3 below shows that, on average, each respondent received COVID-19 information from eight information and communication channels (Mean = 4.86). Many respondents received information through mass media (86.1%) and health workers (72.9%). Further, 68.1% of the respondents received information via social networks, mainly via Facebook and twitter (now called X) while those who received information via government sources comprised 62.3%. Political leaders were the least utilized and trusted source of COVID-19 information (38.7%).

**Table 3:** Risk communication.

| Information source | Yes | No |
| --- | --- | --- |
| Social media | 549(68.1) | 257(31.9) |
| Mass media | 694(86.1) | 112(13.9) |
| Relatives and friends | 422(52.5) | 382(47.5) |
| Health workers | 638(72.9) | 167(20.7) |
| The Internet | 517(64.1) | 289(35.9) |
| Government officials | 502(62.3) | 304(37.7) |
| Political leaders | 312(38.7) | 494(61.3) |
| Print media | 505(62.7) | 301(37.3) |

### Trusted source of information

Figure 1 below shows that trusted information sources were health care professionals (86.7%) The World Health Organization (86.1%), The Center for Disease Control and prevention (CDC – Africa regional Office) (85.5%), Newspapers (81.1%), Local radio and TV news stations (77.3%), Governments websites (75.3%), and family and friends (63.9%), respectively. Facebook (50.6%) and twitter (46.4%) were the least trusted sources of information.

**Figure 1.**
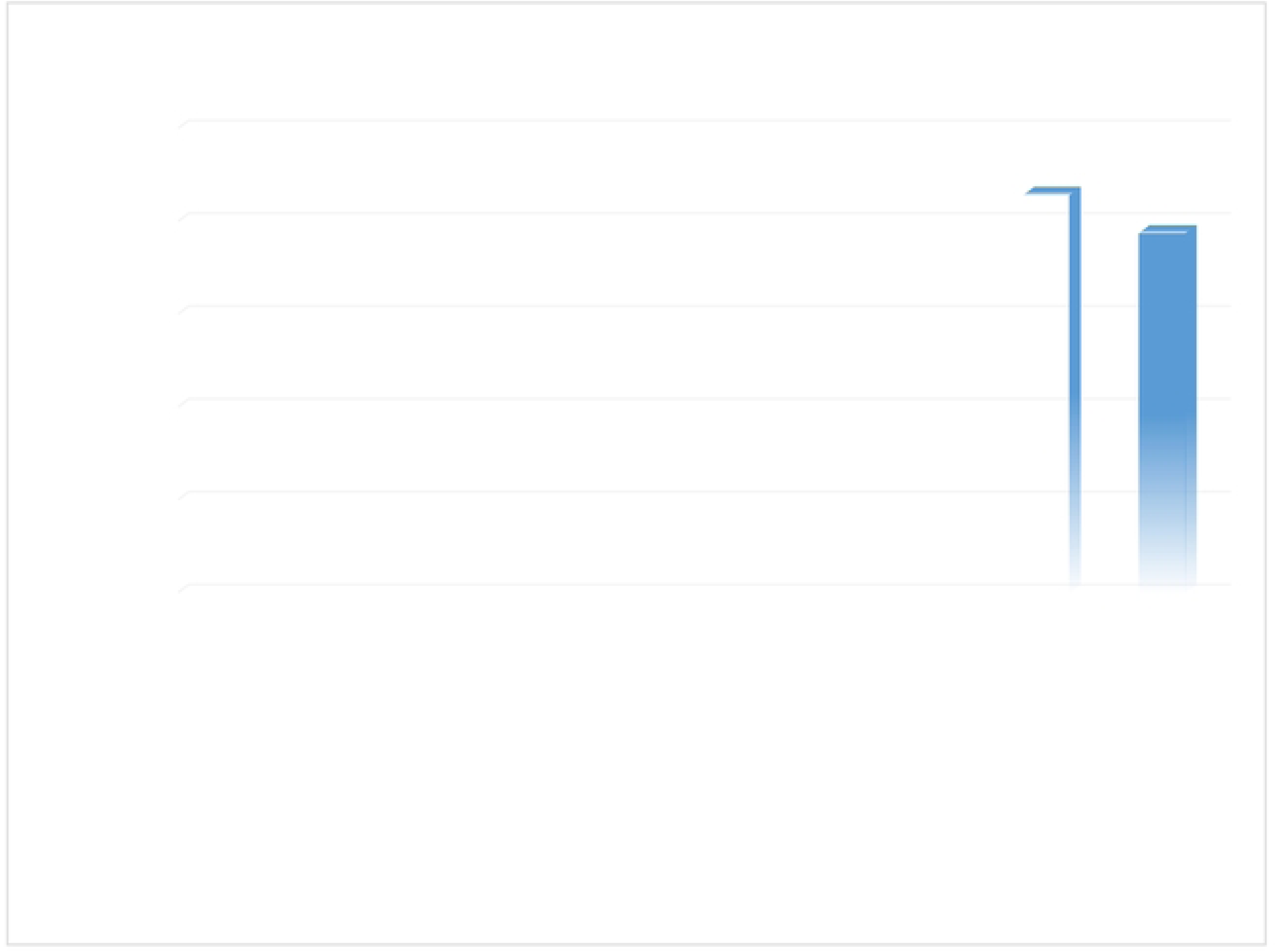
Trusted information source.

### Risk communication by trusted source, its role on risk perception, compliance to COVID-19 mitigation measures and vaccination

Table 4 below shows risk communication by trusted source, its role on risk perception, compliance to COVID-19 mitigation measures and vaccination. Overall, there was significant influence of risk communication on risk perception in terms of risk susceptibility and risk severity (p= 0.001), compliance to covid-19 mitigation measures (p= 0.001), and vaccination acceptance (p= 0.001). Though risk susceptibility, and vaccination rate was low, the results shows that mass media (OR = 1.8, CI = 1.51, 2.18; P= 0.001) and official institutions and professionals (OR = 0.8, CI = 0.75, 0.97; P= 0.001) as trusted sources of information resulted in increased influence on risk susceptibility. Despite low risk susceptibility and vaccination rates, the results indicate that trusted sources of information significantly influenced risk perception. Specifically, exposure to mass media was associated with an increased risk susceptibility (OR = 1.8, CI = 1.51–2.18; P = 0.001), while information from official institutions and professionals was linked to a decreased perception of risk (OR = 0.8, CI = 0.75–0.97; P = 0.001).

**Table 4:**
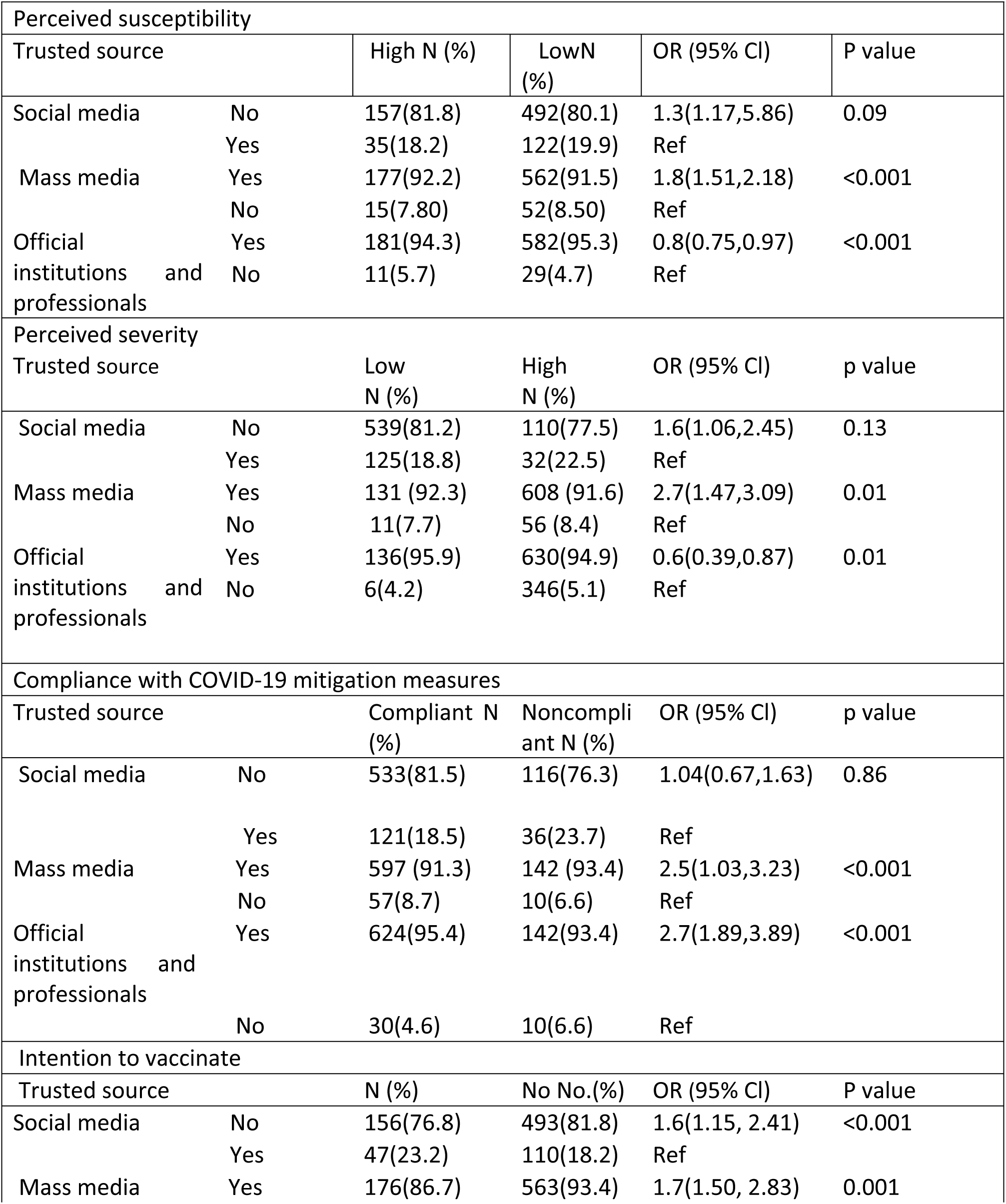
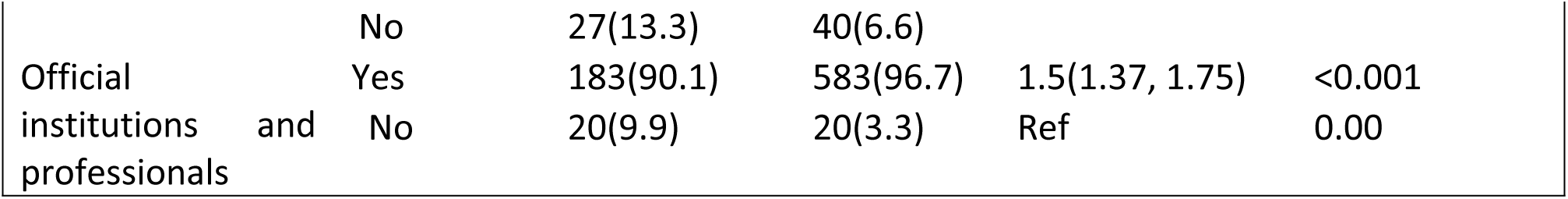
Risk communication and risk perception, compliance to COVID-19 mitigation measures and intention to vaccinate against COVID-19.

**Table 5:**
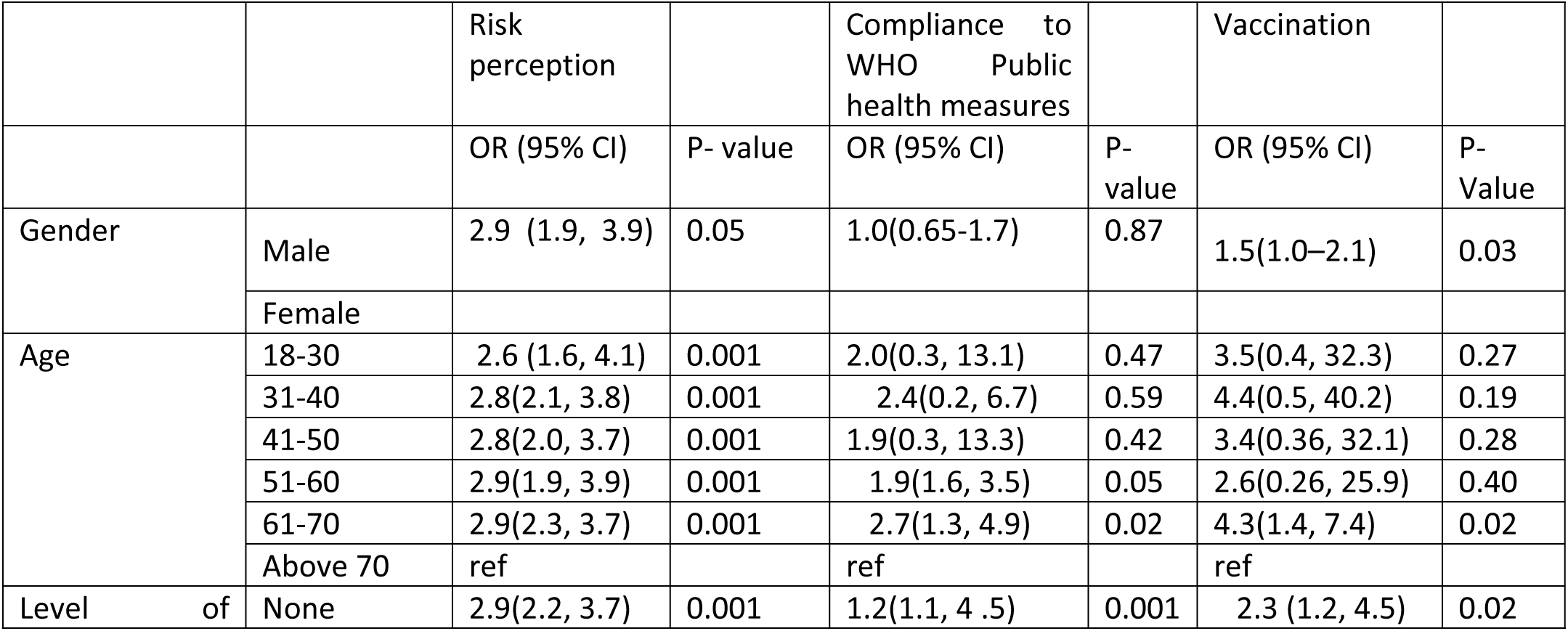
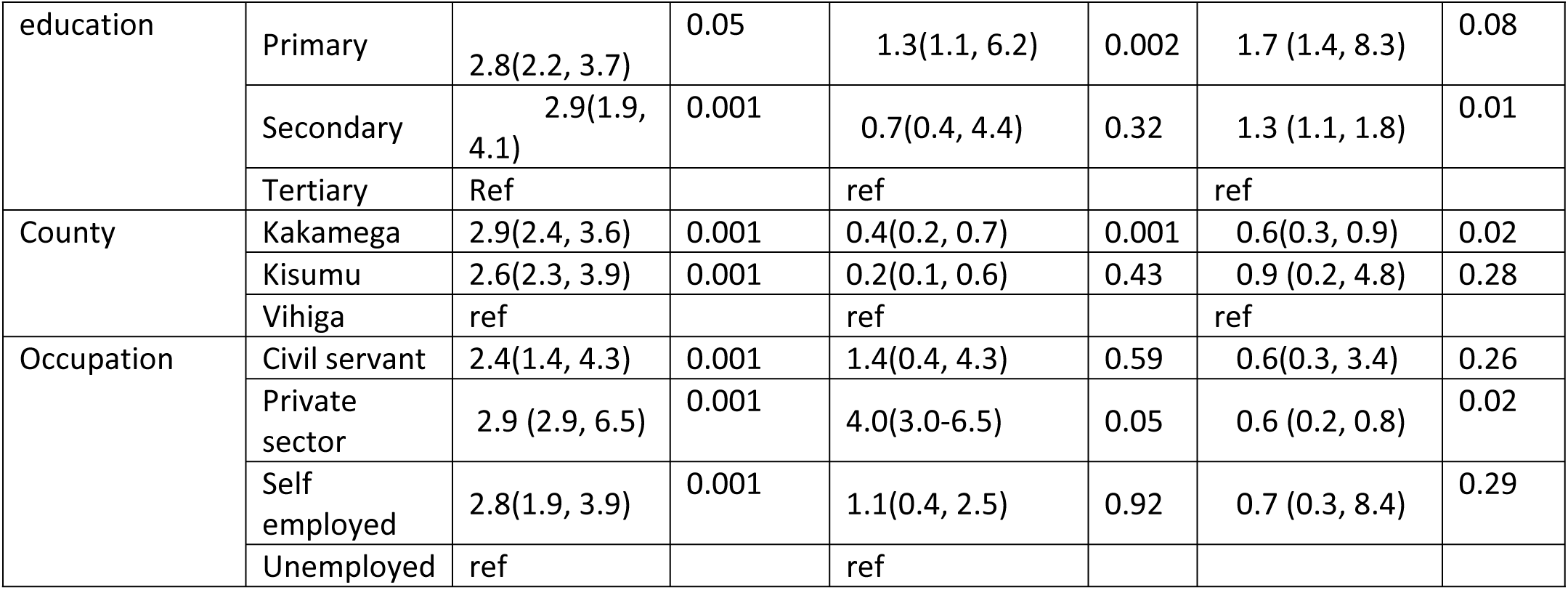
Logistic regression analysis of the association between demographic characteristics and risk perception, Compliance with WHO public health measures and vaccination.

Differences in perceived severity due to mass media and official institutions or professional as trusted sources of communication were significant (P=0.001). The proportion of participants who trusted mass media and official institution and professionals as a source of communication had high perceived severity representing 82.3% and 82.2% respectively compared to only 16.9% who trusted social media as a source of communication. Greater proportions of lower perceived severity were found among participants who trusted social media (79.6%).

Further data reveals that risk communication via official institutions and professionals and mass media raised participants’ perception on risks and compliance to WHO COVID-19 mitigation measures. The proportion of those who complied to the mitigation measures were highest among those who received and trusted information via official institutions and professionals, and mass media compared to social media (81.5% and 80.8% vs. 77.1%). Hence increased odds of 2.7 times and 2.5 times respectively for those who received and trusted information from official institutions and professionals and mass media respectively.

### Logistic regression analysis of the association between demographic characteristics and risk perception, Compliance with WHO public health measures and Vaccination

The study findings revealed that demographic factors played critical role in influencing participants’ perception on risk and eventual public health action(s). Specifically, examination of influence of different demographic factors on risk perception compared to respective reference categories shows that the odds for higher risk perception were significantly increased with higher age groups (50+) (OR = 2.9; 95% CI: 1.9, 3.9), being male (OR =2.9;95% CI: 1.9, 3.9); private sector workers (OR 2.9; 95% Cl; 2.9, 6.5. Further, the odds of significantly higher risk perception were 2.9 times each greater for those who had no education and those who had completed secondary school (OR = 2.9, CI = 2.2, 3.7), (OR = 5.320, CI = 2.9 1.9, 4.1) compared to participants who completed tertiary education respectively.

Strikingly, there was also greater likelihood of participants who were aged between 61-70 years and those who did not attain any level of education, those who completed primary school and those who were working in private sector were more likely to comply to WHO public health measures (OR=2.7, 95% Cl; 1.3, 4.9) and (OR = 1.2, CI = (1.1, 4 .5), (OR=1.3, 95% Cl; 1.1, 6.2) and (OR=4.0, 95% Cl; 3.0-6.5) than those aged above 70 years, participants who completed primary school, and those that were working in private sector respectively. Notably, participants aged 61-70 years, those with no formal education, those who completed primary school, and private sector workers were more likely to comply with WHO public health measures. Specifically, the odds ratios indicated that individuals aged 61-70 had an OR of 2.7 (95% CI: 1.3–4.9), while those with no formal education had an OR of 1.2 (95% CI: 1.1–4.5). Participants who completed primary school had an OR of 1.3 (95% CI: 1.1–6.2), and private sector workers showed the highest likelihood of compliance with an OR of 4.0 (95% CI: 3.0–6.5). These findings suggest a greater likelihood of compliance among these groups compared to individuals over 70 years, those with higher educational attainment, and employees in sectors other than private. Furthermore males (OR = 1.5; Cl= 1.0–2.1; p=0.03); motorcycle riders (OR = 5.22; CI= 3.07-12.67; P=0.03), age between 61-70 years (OR = 4.3; Cl= 1.4, 7.4); p=0.02); and participant who worked in private sector (OR = 0.6; Cl= (0.2, 0.8); p=0.02); had significant association and increased intention for vaccination. Those aged below 60 years, civil servants were not associated with increased intention for vaccination.

## Discussion

This study investigated the role of risk communication and demographic characteristics on risk perception and compliance with WHO public health mitigation measures against COVID-19 in selected rural communities in Western Kenya. Overall compliance with WHO public health mitigation measures against COVID-19 was quite good with 85.3% wearing face mask, 78.9% practiced hand hygiene while 66% avoided hand shaking, hugging and kissing and practiced cough hygiene. These results were comparable to others recorded in the region [36].

Although Social distancing was the least adopted (60.2%) protocol in the current study, the rate was strikingly similar to that reported in parts of Ethiopia (59.2%) suggesting a general phenomenon in the region [37]. One review identified community-level psycho-social and sociological influences (including fear of stigma, misconceptions, lack of trust in authorities, and inconsistencies between personal experience, and information received); and shortcomings in governmental action or communication (including lack of community engagement, poor communication and lack of emotional, financial or material support) as the two leading barriers to adopting social distancing [37]. However, other studies conducted in other African countries cited cultural norms as a leading barrier with some considering the measures anti-social [37, 38, 39].

As regards risk communication, majority of respondents received information through mass media (86.1%) and health workers (72.9%). Furthermore, a large number (68.1%) of the respondents received information via social networks, mainly via Facebook and X (twitter). These results were comparable to those reported by Hailu and colleagues [37], although in their study social media accounted for a higher proportion (80%). A study involving several sub-Saharan Africa (SSA) countries reported similar results across regions, age-groups and gender [40]. Therefore, harnessing the power of social media to influence the quality of information shared with the public on mitigating such severe PHEs is critical especially in defraying misinformation, conspiracy theories and helping establish trust in credible information. This means understanding people’s changing perceptions and attitudes, and the barriers and enablers influencing their ability and motivation to adopt and/or sustain positive health behaviors are critical during and after PHEs.

Additionally, we investigated respondents’ confidence regarding three main categories of communication media comprising social media, mass media, official institutions, and professionals. The findings showed that social media (including Facebook (50.6%) and X (twitter) (46.4%)) were the least-trusted sources of information. These proportions were not surprisingly quite high considering the infodemic around COVID-19. That half the respondents trusted COVID-9 related information on social media put a large proportion at risk of misinformation [41,42].

This study found that demographic factors influenced participants’ perception on risk. The odds for risk perception were significantly increased with higher age groups (above 50) (OR = 2.9; 95% CI: 1.9, 3.9) and being male (OR =2.9;95% CI: 1.9, 3.9). Studies involving other sub-Saharan Africa countries have reported similar results regarding relationship between age and risk perception [31]. This observation is explained by public awareness regarding older individuals having significantly higher COVID-19 related severe disease and deaths than young individuals. Our findings regarding male participants having higher odds of risk perception, however, was different from that observed in some other SSA countries where risk perception did not vary significantly with sex [43].

Interestingly, the odds of risk perception were 2.9 times greater for participants with no formal education (OR = 2.9, CI = 2.2–3.7) and 5.32 times greater for those who completed secondary school (OR = 5.32, CI = 2.9–4.1) compared to those with tertiary education. This finding is unexpected, as higher educational attainment typically correlates with a better understanding of COVID-19-related risks. However, our results align with a study conducted across seven sub-Saharan African countries, which found that respondents with primary education had a higher perception of risk than those with secondary and tertiary education [44]. Furthermore, this study found greater likelihood of compliance with WHO public health mitigation measures among participants who were older and with lower educational attainment. This observation could have been as a result of their higher risk perception which mediated compliance. This was in concurrence with the findings of Asnakew and colleagues [26] in the Federal Democratic Republic of Ethiopia. However, our finding that there was no statistically significant difference in gender as regards to compliance was contrary to the findings of Asnakew and others [36] who reported females were more likely to have better compliance with preventive behavior than their male counterparts.

Moreover, this study found that risk communication particularly via trusted media such as mass media increased the odds of both risk perception and preventive behaviors’ compliance. The proportion of those who complied with COVID-19 mitigation measures were highest among those who received and trusted information through mass media (80.8%) resulting in increased odds of 2.5 times. Our findings concurred with the findings of a study conducted in parts of Nigeria where 60% of respondents indicated they were persuaded by mass media messaging to comply with COVID-19 public health measures including wearing facemasks [45]. Other studies confirmed that exposure to risk communication through mass media improves public compliance directly and indirectly through the mediating roles of public understanding and risk perception [46, 47]. Therefore, vaccine education campaigns with accurate, credible, and transparent information on the efficacy and safety of the vaccines, among other things, should be communicated to the public in a timely manner to ensure effective all-inclusive immunization [48].

The findings of this study bolster the evidence that mass media is one of the strategies that provides powerful tools and platforms for risk communication that can greatly influence public perceptions of the issues of societal concern and enhance compliance with public health protocols and related measures. These can be harnessed for expanded reach and engagement, targeted messages in public health campaigns, peer influence, and increased community networks’ support and participation in driving desired behavior changes.

## Strengths and Limitations

### Strengths

The present study benefits from a large and diverse sample, which is more representative of the general population across different communities and counties in Kenya, thereby enhancing the generalizability of the findings. The study employed a priori power analysis to determine the sample size, ensuring sufficient statistical power to detect meaningful effect sizes before the study began. This methodological rigor strengthens the reliability of the results. Additionally, while Kenya’s COVID-19 measures generally align with WHO recommendations, variations in regional policies and public health campaigns may affect compliance differently across areas. This diversity in policy implementation provides a rich context for understanding the complexities of noncompliance and its regional variations.

### Limitations

However, there are several limitations to consider. First, the sample was predominantly drawn from western Kenya, with a younger age distribution and a slight male predominance. This regional and demographic skew may limit the broader applicability of the findings. Second, the cross-sectional design of the study presents a limitation, as it only captures data at one point in time, preventing conclusions about causality or the evolution of behaviors throughout the pandemic. The potential influence of unobserved confounding variables over time means that the statistical associations observed may not reflect causal relationships. Finally, the reliance on self-reported compliance behaviors introduces the risk of response bias, as participants may underreport or overreport their behaviors. This introduces the possibility that the self-reported data may not accurately reflect participants’ actual actions, as social desirability or memory recall biases could affect the honesty and precision of responses.

While the study provides valuable insights into the role of risk communication and demographic factors on public health emergency responses during the pandemic, caution is needed when interpreting the results due to the limitations inherent in the sampling, design, and measurement approaches used.

## Conclusions

The study reveals that a majority of the participants effectively adhered to one or more WHO preventive measures to avoid COVID-19 infection. The primary source of trusted information for participants was television, followed by newspapers. Risk communication through official institutions, professionals, and mass media enhanced participants’ risk perception and compliance with COVID-19 mitigation measures. Demographic factors significantly influenced participants’ risk perception and subsequent public health actions. Higher risk perception was notably associated with older age groups (above 50 years), male private sector workers, and individuals with higher education levels. In contrast, younger individuals and those with lower education levels exhibited lower adoption of preventive measures.

Evidence suggests that increased risk perception correlates with heightened preventive behavior among the general population. Therefore, targeted risk communication and awareness campaigns are essential to bolster desired public health practices, particularly among demographic segments with lower adoption rates. Focusing on younger individuals and those with less education could improve overall adherence to preventive measures and ensure sustained public health practices throughout the COVID-19 pandemic while supporting those who were compliant to sustain the behaviors. Governments and health organizations ought to prioritize investment in rural communication infrastructure, through continuous engagement with communities to build trust and ensure consistent and accurate dissemination of information, and monitoring and evaluation of communication strategies to identify and address gaps in real time. This means understanding people’s changing perceptions and attitudes, and the barriers and enablers influencing their ability and motivation to adopt and/or sustain positive health behaviors are critical during and after PHEs. Effective coordinated risk communication in rural communities is a dynamic, iterative process, requiring multi-sectoral and multi-dimensional approaches considerate of contextual factors and active community engagement.

## Acknowledgments

We are grateful to all the research assistants and field coordinators who helped with the study. We acknowledge participation of those who accepted to be interviewed for their time and shared experiences with us.

## Funding Statement

This research received no external funding.

## Author Contributions

W.C.—conceptualized the study, methods, data analysis, and reporting and manuscript revision; F.J.N.— conceptualized the study and significantly worked on the manuscript, revised and provided approval for submission. C.K.N.—conceptualized the study, data collection, manuscript preparation and revisions. W.A. — manuscript preparation and revisions. All authors have read and agreed to the published version of the manuscript.

## Institutional Review Board Statement

The study was conducted according to the guidelines of the Declaration of Helsinki and approved by the Ethics Committee of University of Eastern Africa, Baraton. Approval Code: (UEAB/REC/50/03/2021).

## Informed Consent Statement

Informed consent was obtained from all subjects involved in the study by signing a consent form which included an explanation about voluntary participation and the purpose of the study.

## Data Availability Statement

The datasets used and/or analyzed during the current study are contained within this article. Any additional data or clarification available upon reasonable request.

## Conflicts of Interest

The authors declare no conflict of interest.

## Notes

### Competing Interest Statement

The authors have declared no competing interest.

### Author Declarations

The approval and permission to conduct the study were obtained from the University of Eastern Africa, Baraton Institutional Research Ethics Committee (IREC), and the National Commission for Science, Technology and Innovation (NACOSTI) in Nairobi, respectively.

